# Seroepidemiological and genomic investigation of COVID-19 spread in North East region of India

**DOI:** 10.1101/2022.01.25.22269702

**Authors:** Romi Wahengbam, Pankaj Bharali, Prasenjit Manna, Tridip Phukan, Moirangthem Goutam Singh, Gayatri Gogoi, Yasmin Begam Tapadar, Anil Kumar Singh, Rituraj Konwar, Channakeshavaiah Chikkaputtaiah, Natarajan Velmurugan, Selvaraman Nagamani, Hridoy Jyoti Mahanta, Himakshi Sarma, Ravi Kumar Sahu, Prachurjya Dutta, Sawlang Borsingh Wann, Jatin Kalita, G Narahari Sastry

**Affiliations:** Centre for Infectious Diseases, CSIR–North East Institute of Science and Technology, Jorhat, Assam, India; Biological Sciences and Technology Division, CSIR–North East Institute of Science and Technology, Jorhat, Assam, India; Branch Laboratory, CSIR–North East Institute of Science and Technology, Naharlagun, Papumpare, Arunachal Pradesh, India; Advanced Computation and Data Sciences Division, CSIR– North East Institute of Science and Technology, Jorhat, Assam, India; Academy of Scientific and Innovative Research (AcSIR), Ghaziabad, India

**Keywords:** SARS-CoV-2, virus, test positivity rate, seroprevalence, neutralizing antibody, whole genome sequencing, vaccination

## Abstract

Seroepidemiology and genomics are valuable tools to investigate the transmission of COVID-19. We utilized qRT-PCR, serum antibody immunoassays, and whole genome sequencing to examine the spread of SARS-CoV-2 infections in North East (NE) region of India during the first and second pandemic waves (June 2020 to September 2021). qRT-PCR analysis was performed on a selected population from NE India during June 2020 to July 2021, and metadata were collected for the region. Seroprevalence and neutralizing antibody immunoassay were studied on selected individuals (n=2026) at three time points (August 2020, February 2021 and June 2021), as well as in a cohort (n=35) for a year (August 2020 to August 2021). SARS-CoV-2 genomes of 914 qRT-PCR positive samples (June 2020 to September 2021) were sequenced and assembled, and those obtained from the sequence databases were analyzed. Test positivity rates in first and second waves were 6.34% and 6.64% in the state of Assam, respectively, and a similar pattern was observed in other NE states. Seropositivity in August 2020, February 2021, and June 2021 were 10.63%, 40.3% and 46.33% respectively, and neutralizing antibody prevalence were 90.91%, 52.14%, and 69.30% respectively. The cohort group showed the presence of stable neutralizing antibody throughout the year. Normal variants dominated the first wave, while the variant of concerns (VOCs) B.1.617.2 and AY-sublineages dominated the second wave, and identified mostly among vaccinated individuals. All eight states of NE India reported numerous incidences of SARS-CoV-2 VOCs, especially B.1.617.2 and AY sublineages, and their prevalence co-related well with high TPR and seropositivity rate in the region. High infection and seroprevalence of COVID-19 in NE India during the second wave was associated with the emergence of VOCs. Natural infection prior to vaccination provided higher neutralizing activity than vaccination alone.

## Introduction

North East (NE) India is a unique region with eight states and strategically important as a gateway for South East Asia to the rest of India. NE India is sharing 99% international border with Myanmar, China, Bhutan, Nepal and Bangladesh. This region is crucial in the India’s effort on tracing, testing, isolation and treating of COVID-19. Among NE India, the state of Assam, Tripura, Meghalaya, and Nagaland showed high COVID-19 incidences, and these states constitute almost 76.93% of the NE India population. Strict surveillance of SARS-CoV-2 in the region is important to restrict transnational SARS-CoV-2 transmission.

Seroepidemiological studies are useful to understand the true burden and transmission of SARS-CoV-2. Till now, only a limited number of seroepidemiological studies of SARS-CoV-2 have been reported in India [1]. Recently, genomic surveillance of SARS-CoV-2 helped to understand the spread and evolution of the virus particularly variants of concern (VOC) and variants of interest (VOI) [2-4]. As per the Indian SARS CoV-2 Genomics Consortium (INSACOG), only 2.36% genome sequencing reported from NE India [5]. This clearly indicates a critical dearth of genome surveillance in this region.

This study investigates on COVID-19 infectivity, seroepidemiology and genome surveillance during first wave (June-September 2020), intermittent phase (February 2021) and second wave (May-September 2021) in various states of NE India. Furthermore, a seroepidemiological cohort study was conducted for a year to assess the presence of different antibodies and their neutralizing efficacy against SARS-CoV-2 in pre-and post-vaccination.

## Methods

### SARS-CoV-2 qRT-PCR testing

Naso-oropharyngeal swab samples (n=35255) during first wave (June-September 2020) and second wave (May-July 2021) were collected and tested using Indian Council of Medical Research (ICMR), New Delhi approved qRT-PCR method.

### Immunoassays

Participants were enrolled in August 2020 (n=724), February 2021 (n=866), and June 2021 (n=436) for the present study (Supplementary Figure S1). A total of 2026 participants were interviewed with a questionnaire on the demographic features, anthropometric parameters and histories of clinical features, COVID-19 symptoms and infection, hospital visits and exposure to COVID-19 patients. Serums obtained from 5 mL venous blood that were collected from each participant were subjected to IgG-N (nucleocapsid), IgG-S (spike protein), IgM-S (spike protein) and neutralizing antibody immunoassays specific to SARS-CoV-2. IgG-N antibodies were measured using Elecsys Anti-SARS-CoV-2 kit (Roche Diagnostics, Germany) as per manufacturer’s protocol. IgG-S and IgM-S antibodies were detected using VoxPress New Corona Virus (COVID-19) IgG / IgM Rapid Test kit (Voxtur Bio Ltd, India) following manufacturer’s instructions. Samples that were seropositive for IgG-N, IgG-S and/or IgM-S were further tested for neutralizing antibody response against spike protein using GENScriptcPass SARS-CoV-2 Neutralization Antibody Detection Kit (GenScript, USA), according to the manufacturer’s protocol. A cohort group of 35 individuals were also followed up for a year to monitor SARS-CoV-2 antibodies and their neutralizing efficacy against pre-and post-vaccination at multiple time points (10^th^ August 2020, 10^th^ February 2021, 10^th^ June 2021, 10^th^ July 2021, 16^th^ July 2021, 24^th^ July 2021, 31^st^ July 2021 and 14^th^ August 2021).

### SARS-CoV-2 genome sequencing

SARS-CoV-2 whole genome sequencing was performed using the LoCost ARTIC nCoV-2019 multiplex PCR tiling amplicon sequencing approach [6, 7]. Nine hundred and fourteen qRT-PCR positive samples from first wave (July-August 2020) and second wave (May-September 2021) were sequenced on a MinION Mk1c sequencer (Oxford Nanopore Technologies) using NEB Next ARTIC SARS-CoV-2 Companion Kit (New England Biolabs, UK), Native Barcoding Expansion Kit (EXP-NBD196) and Ligation Sequencing Kit (SQK-LSK-109). Each sequencing run had a no-template negative control from PCR amplification step. The raw FAST5 data were basecalled and demultiplexed using MinION Mk1C MinKNOW (version 21.05.8). The quality-filtered demultiplexed FASTQ reads were aligned with Wuhan-Hu-1 reference SARS-CoV-2 genome (MN908947.3) followed by variant calling and consensus genome assembly using ARTIC nCoV-2019 bioinformatics pipeline v1.1.0 (https://artic.network/ncov-2019/ncov2019-bioinformatics-sop.html) implemented at EPI2MELabs Jupyter notebook server v1.1.2 (Oxford Nanopore Technologies). The sequencing depth was capped at 100X coverage, and N bases masked the positions with a depth lower than 100 X. Mutation calling and assignment of clades and lineages to the genomes were carried out using NextStrain’s NextClade v.154 (https://clades.nextstrain.org/) and Pangolin COVID-19 Lineage Assigner v3.1.16. Variant calling datasets and genome sequences for the NE states of India and its neighboring countries were also retrieved from the data hub of INSACOG (http://clingen.igib.res.in/covid19genomes/), and the GISAID’s EpiCoV database (https://www.gisaid.org/). The sequence alignment files of the genomes were used to construct maximum-likelihood phylogenetic tree using IQ-TREE server (version 1.6.12) and visualized in Fig Tree version 1.4.4. The whole genome sequences generated in this study were deposited to GISAID’s EpiCoV database.

### Ethical statement

All subjects participated on a voluntarily basis and written informed consent was obtained from each participants. The study was approved by the Institutional Human Ethics Committee of CSIR-NEIST (IHEC/NEIST20-21/201).

## Results

### COVID-19 incidences during first and second waves

In March 2020, the first SARS-COV-2 infected case was reported from NE India [8]. Subsequently, COVID-19 incidences gradually spread throughout the NE states. The first COVID-19 infection reported in Assam on 31^st^ March 2020 and registered highest daily record of 2,354 COVID-19 positive cases on 18^th^ August 2020 with total tally of 82,201 infections and 203 deaths (Figure 1). The incidence of second wave was first observed from the beginning of April 2020 and the highest daily registered cases were 5704 on 27^th^ May 2021 in Assam. The SARS-CoV-2 test positivity rate (TPR) in Assam during first wave (June-September 2020) and second wave (May 2021 -July 2021) was 6.34% and 6.64%, respectively. The outbreak of second wave was also considerably high in other nearby states like Manipur, Meghalaya, Mizoram and Sikkim (Figure 1).

**Figure 1:**
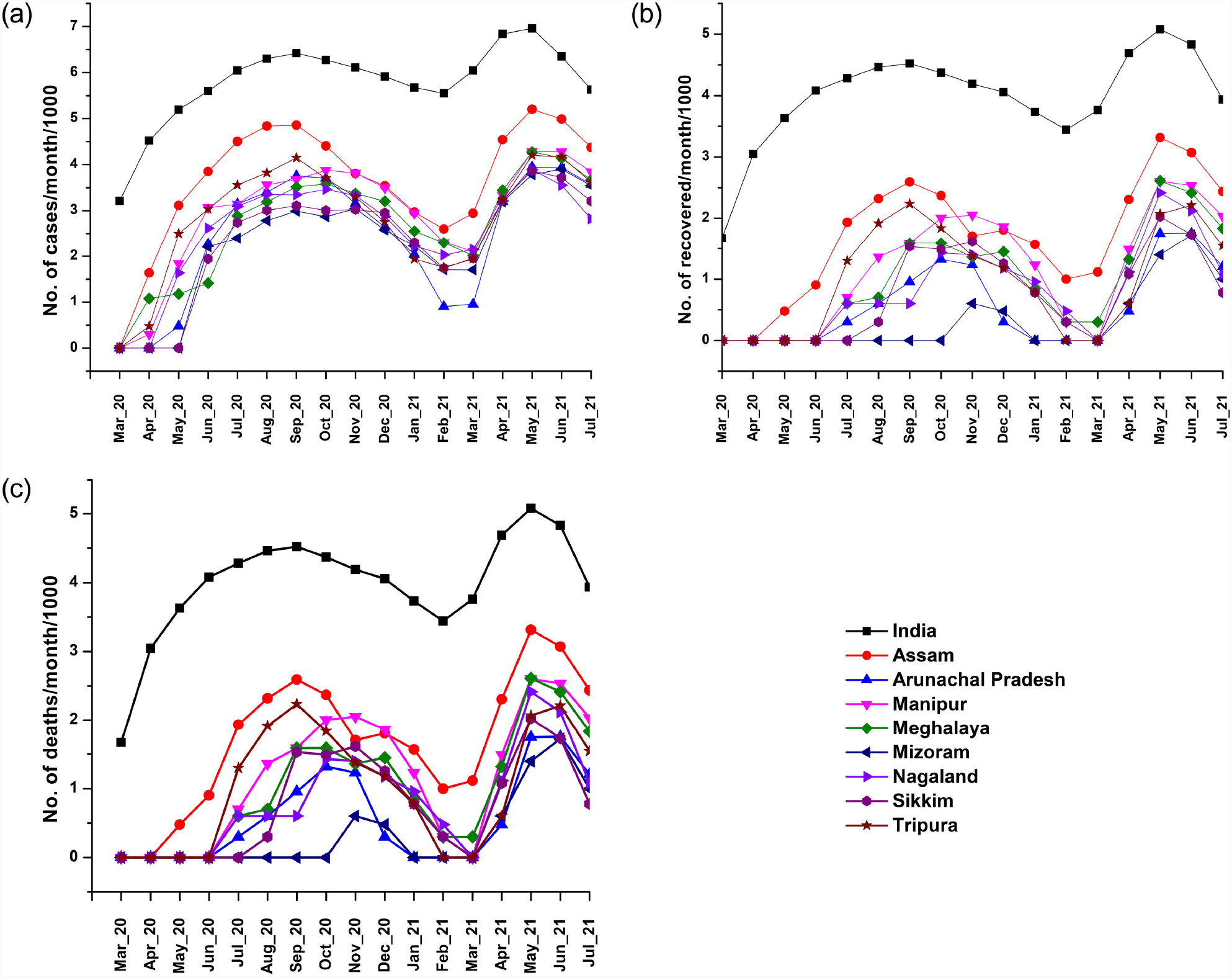
Trend of COVID-19 (a) positivity (b) recovery (c) death in India and states of North East India during first wave and second wave (April 2020 – July 2021).

During first wave (June-September 2020), out of 12879 samples tested from Jorhat district, Assam, 571 tested positive for COVID-19 (TPR: 4.43%). However, in second wave (May-July 2021), out of 3416 samples, 248 tested positive for COVID-19 (TPR: 7.26%). The TPR increased with age in first wave, though it was not statistically significant but observed higher infection in older age group. However, in second wave, a strong negative correlation was observed between TPR and age indicating higher infection in younger subjects (Supplementary Table S1).

### Seroprevalence during first and second waves

Immunoassays for serum IgG-N, IgG-S, IgM-S and neutralizing antibodies were conducted at three-different time points during August 2020, February 2021 and June 2021 with established cohorts of 724, 866, and 436 subjects (n=2026) (Table 1). The anti-SARS-CoV-2 IgG-N, IgG-S, and IgG-M antibodies were found to be 10.63%, 8.83%, and 1%, respectively, during the first wave, while the antibody prevalence in the intermittent phase and second wave were 40.3%, 16.85%, and 3.49%, and 46.33%, 15.82%, and 3.66%, respectively. The frequencies of neutralizing antibody for the above-mentioned periods were found to be 90.91%, 52.14%, and 69.30%, respectively. The prevalence of seropositivity increased while the frequency of neutralizing antibodies decreased both in the intermittent phase and second wave compared to those observed in the first wave (Supplementary Table S2, Table S3 & Table S4). The prevalence of seropositivity and neutralizing antibody decreased with age groups in first wave indicating higher infection in older subjects while in contrast second wave showed an increase in seropositivity and neutralizing antibody with age indicating higher infection in younger subjects.

**Table 1:**
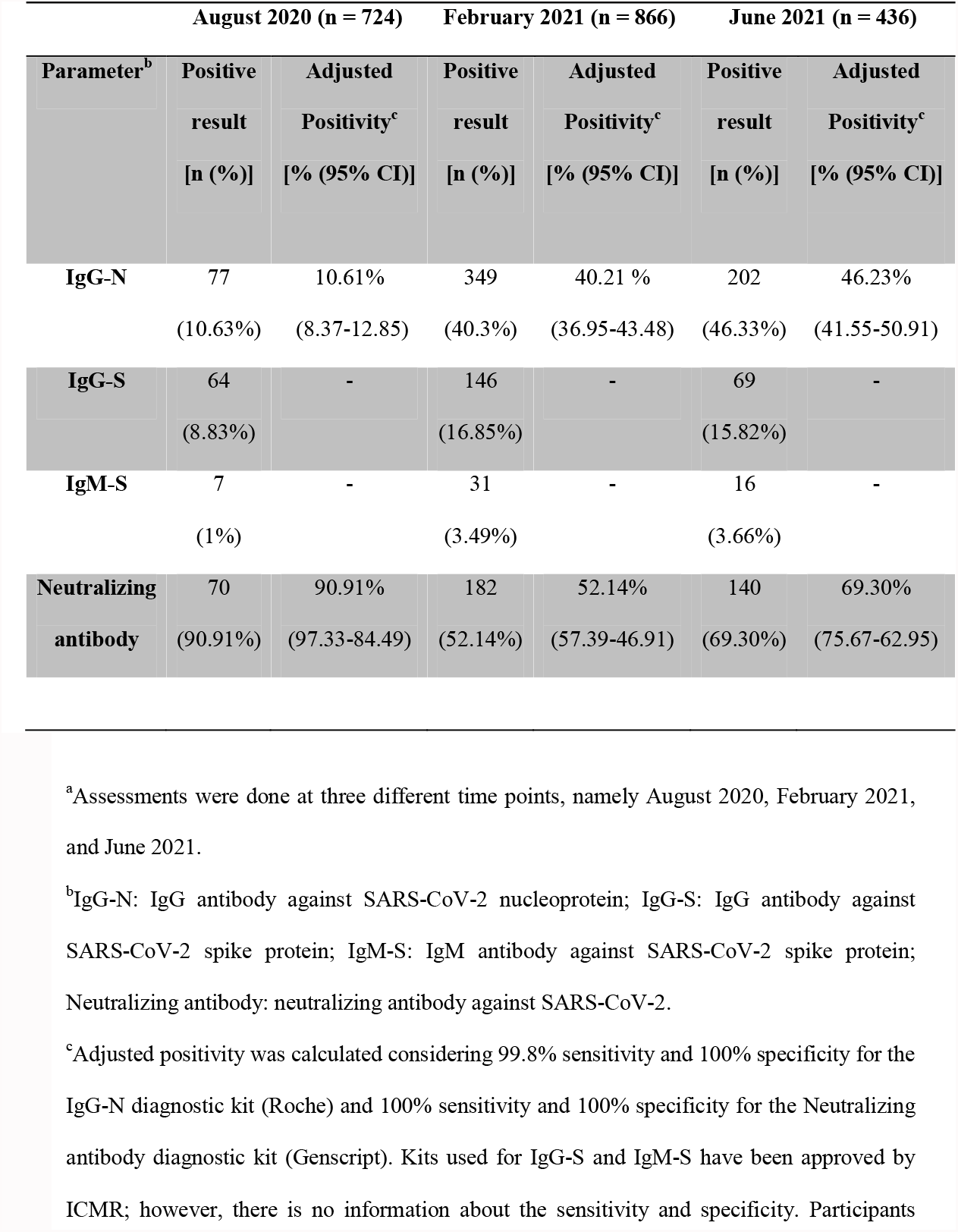
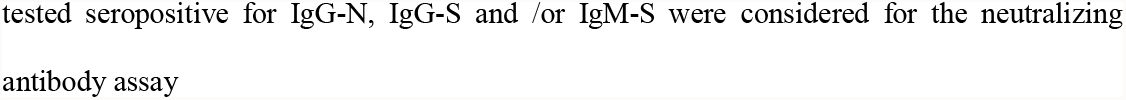
Seropositivity of the participants against SARS-CoV-2^a^.

|  | August 2020 (n = 724) |  | February 2021 (n = 866) |  | June 2021 (n = 436) |  |
| --- | --- | --- | --- | --- | --- | --- |
| Parameter<sup>b</sup> | Positive | Adjusted | Positive | Adjusted | Positive | Adjusted |
|  | result | Positivity<sup>c</sup> | result | Positivity<sup>c</sup> | result | Positivity<sup>c</sup> |
|  | [n (%)] | [% (95% CI)] | [n (%)] | [% (95% CI)] | [n (%)] | [% (95% CI)] |
| **IgG-N** | 77 | 10.61% | 349 | 40.21 % | 202 | 46.23% |
|  | (10.63%) | (8.37-12.85) | (40.3%) | (36.95-43.48) | (46.33%) | (41.55-50.91) |
| **IgG-S** | 64 | - | 146 | - | 69 | - |
|  | (8.83%) |  | (16.85%) |  | (15.82%) |  |
| **IgM-S** | 7 | - | 31 | - | 16 | - |
|  | (1%) |  | (3.49%) |  | (3.66%) |  |
| **Neutralizing** | 70 | 90.91% | 182 | 52.14% | 140 | 69.30% |
| **antibody** | (90.91%) | (97.33-84.49) | (52.14%) | (57.39-46.91) | (69.30%) | (75.67-62.95) |

<sup>a</sup>Assessments were done at three different time points, namely August 2020, February 2021, and June 2021.
<sup>b</sup>IgG-N: IgG antibody against SARS-CoV-2 nucleoprotein; IgG-S: IgG antibody against SARS-CoV-2 spike protein; IgM-S: IgM antibody against SARS-CoV-2 spike protein; Neutralizing antibody: neutralizing antibody against SARS-CoV-2.
<sup>c</sup>Adjusted positivity was calculated considering 99.8% sensitivity and 100% specificity for the IgG-N diagnostic kit (Roche) and 100% sensitivity and 100% specificity for the Neutralizing antibody diagnostic kit (Genscript). Kits used for IgG-S and IgM-S have been approved by ICMR; however, there is no information about the sensitivity and specificity. Participants
tested seropositive for IgG-N, IgG-S and /or IgM-S were considered for the neutralizing antibody assay

### Emergence of SARS-CoV-2 variants during first and second wave

SARS-CoV-2 genomes of 914 qRT-PCR positive samples collected from Assam and Arunachal Pradesh during first wave (591 samples) (July–November 2020) and second wave (323 samples) (May – September 2021) were sequenced and assembled. Out of total 815 samples that passed the QC read filtering and lineage assignment, overall 39 variants were detected, including VOC B.1.617.2 (Delta) (n=158), AY sublineages (n=112), and VOI B.1.617.1 (Kappa) (n=1). Thirty two normal variants were detected in first wave predominated by B.1.36 (n=329), B.1.1.201 (n=105) and B.1 (n=36), while second wave was identified with 9 variants predominated by VOC B.1.617.2 (Delta) (n=157), AY sublineages (n=112) and VOI B.1.617.1 (Kappa) (n=1). Among 162 samples that had vaccination details, VOCs and VOI were identified mostly in vaccinated individuals (n=94, 58.02%), among which the highest infections occurred in single-dose vaccinated individuals (n=51, 54.26%), followed by double-dose vaccine breakthrough-infection (n=38, 40.43%). The maximum-likelihood time-resolved phylogenetic tree with distribution of the detected variants is showed in Figure 2.

**Figure 2.**
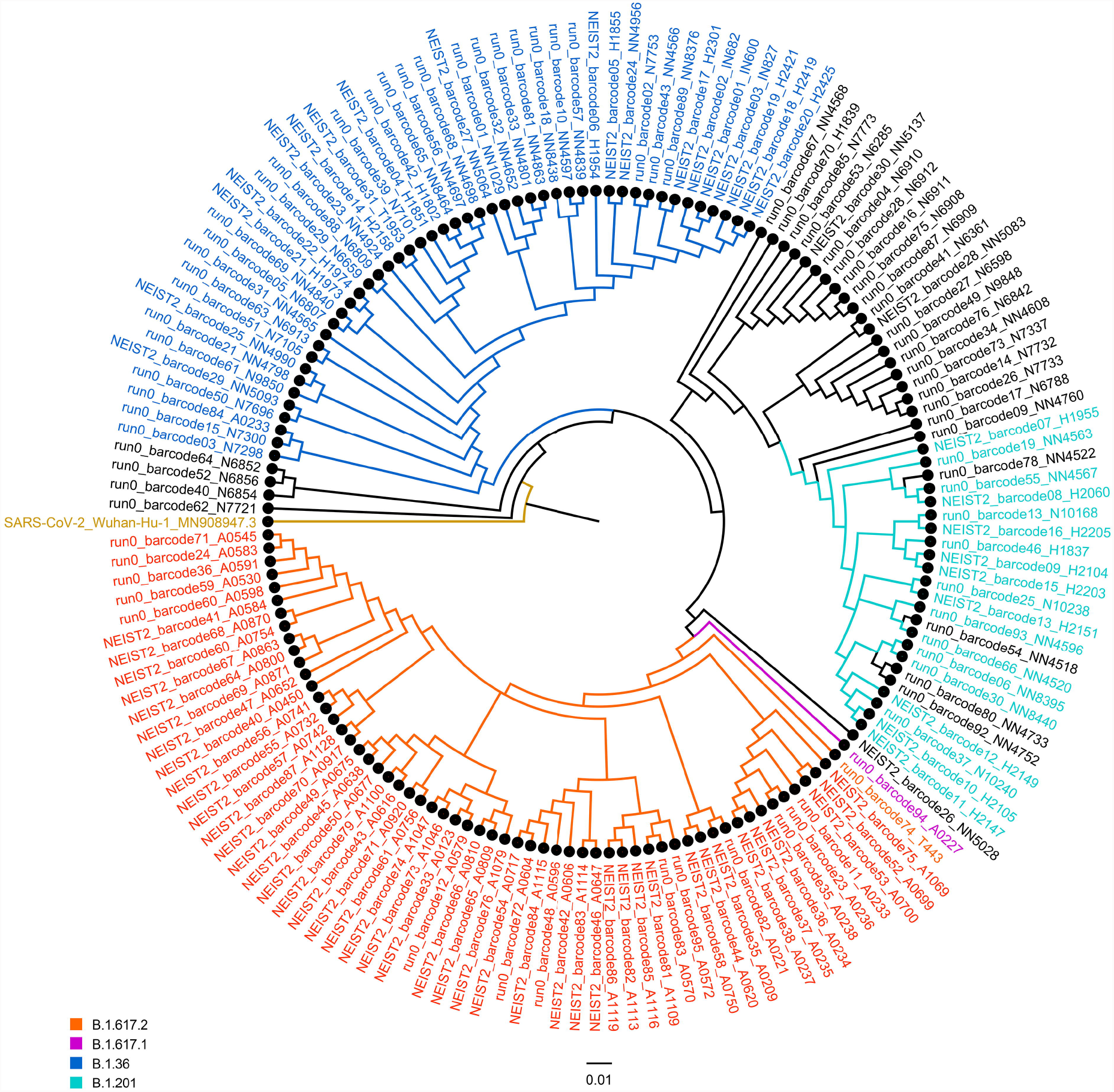
Maximum likelihood phylogenetic tree based on the sequences of SARS-CoV-2 whole genomes from Assam showing distribution of the variants rooted with reference to the Wuhan-Hu-1 reference genome (MN908947.3). The tree was constructed using the IQ-TREE web server implemented using IQ-TREE multicore version 1.6.12. The best-fit model was chosen using Bayesian Information Criterion using the default parameters. The best-fit model used was GTR+F+I with 1000 bootstrap replicates to generate the consensus tree. The tree file generated was imported to FigTree version 1.4.4 and annotated and visualized to generate the time-resolved tree.

The overall number of mutations in different genes across the variants are represented in Supplementary Figure S2 & Figure S3. Total 3376 mutations (synonymous: 1208; missense: 2149; non-sense: 19) were observed across the SARS-CoV-2 genomes, wherein 865 mutations (synonymous: 451; missense: 395; nonsense: 19) were exclusively observed in genomes from first wave, while 1731 mutations (synonymous: 447; missense: 1284) were observed exclusively in genomes from second wave (Supplementary Table S5). Highest mutation frequencies were predominantly observed in ORF1a, ORF1b, S and N genes (Table 2). An increase in the frequency of mutation in second wave was observed in S, ORF3a, ORF7a, ORF7b and 3’ UTR of the genomes. The mutation pattern and frequency among the variants shifted between the two waves, and this variation was predominantly associated with non-synonymous mutations leading to increase in amino acid changes in S, N, ORF1a, ORF1b, ORF3a, ORF7a and ORF7b during the second wave as compared to the first wave. Interestingly, S-gene initially acquired three non-sense mutations, namely C27972T (Q27*) (15.79%), G28221T (E110*) (3.16%), and G28083T (E64*) (1.05%) during first wave, which was subsequently lost in second wave (Supplementary Table S5). Moreover, deletion mutations were predominantly observed only in second wave among the Delta variants in S, ORF3a and ORF8 genes. The deletions in S and ORF8 genes were observed across all Delta genomes analyzed, while the deletions in ORF3a were detected only in three Delta genomes. The C>T transition (43.1%) and G>T transversion (25.3%) were dominant among the overall mutations (Supplementary Table S5).

**Table 2:**
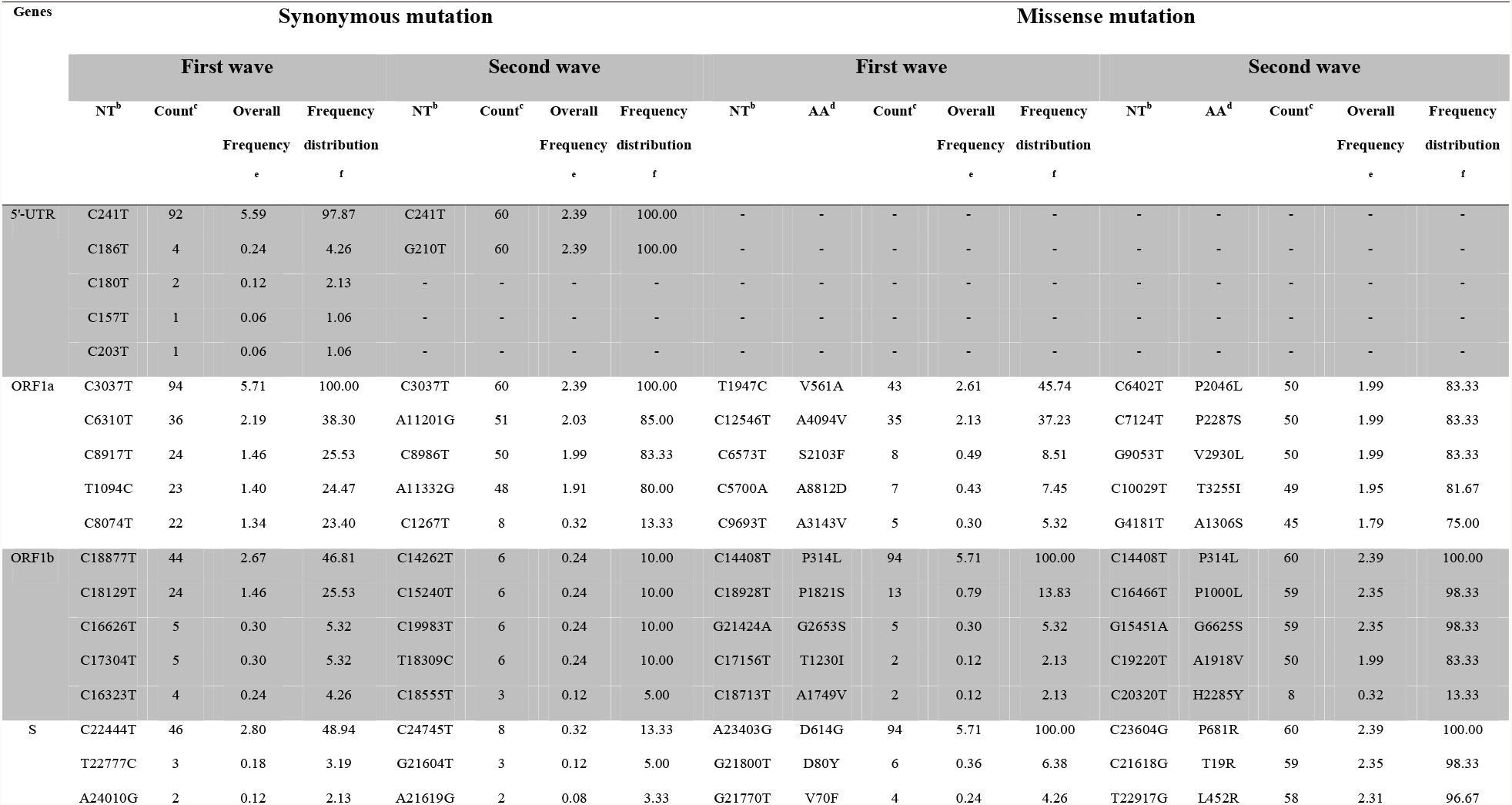
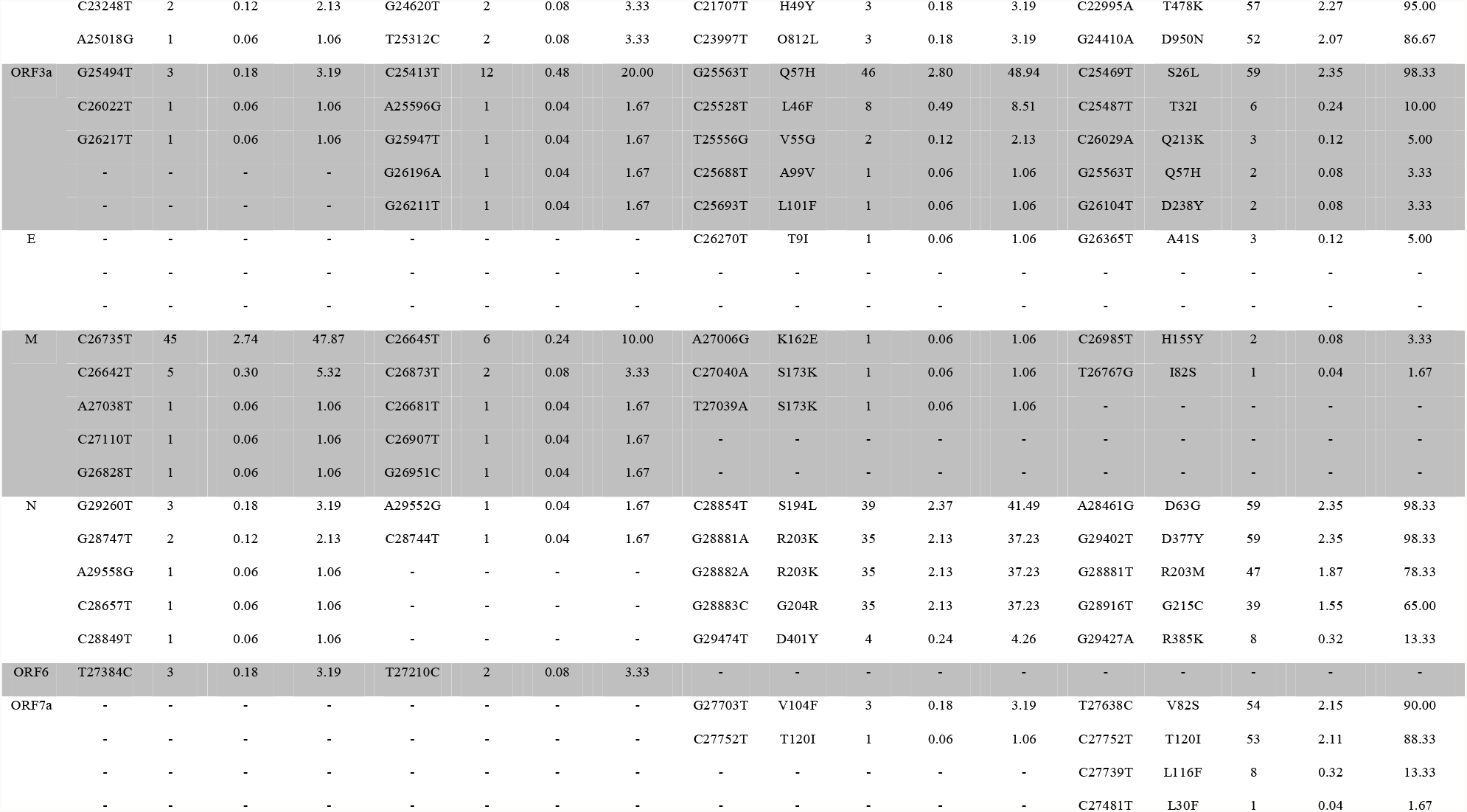
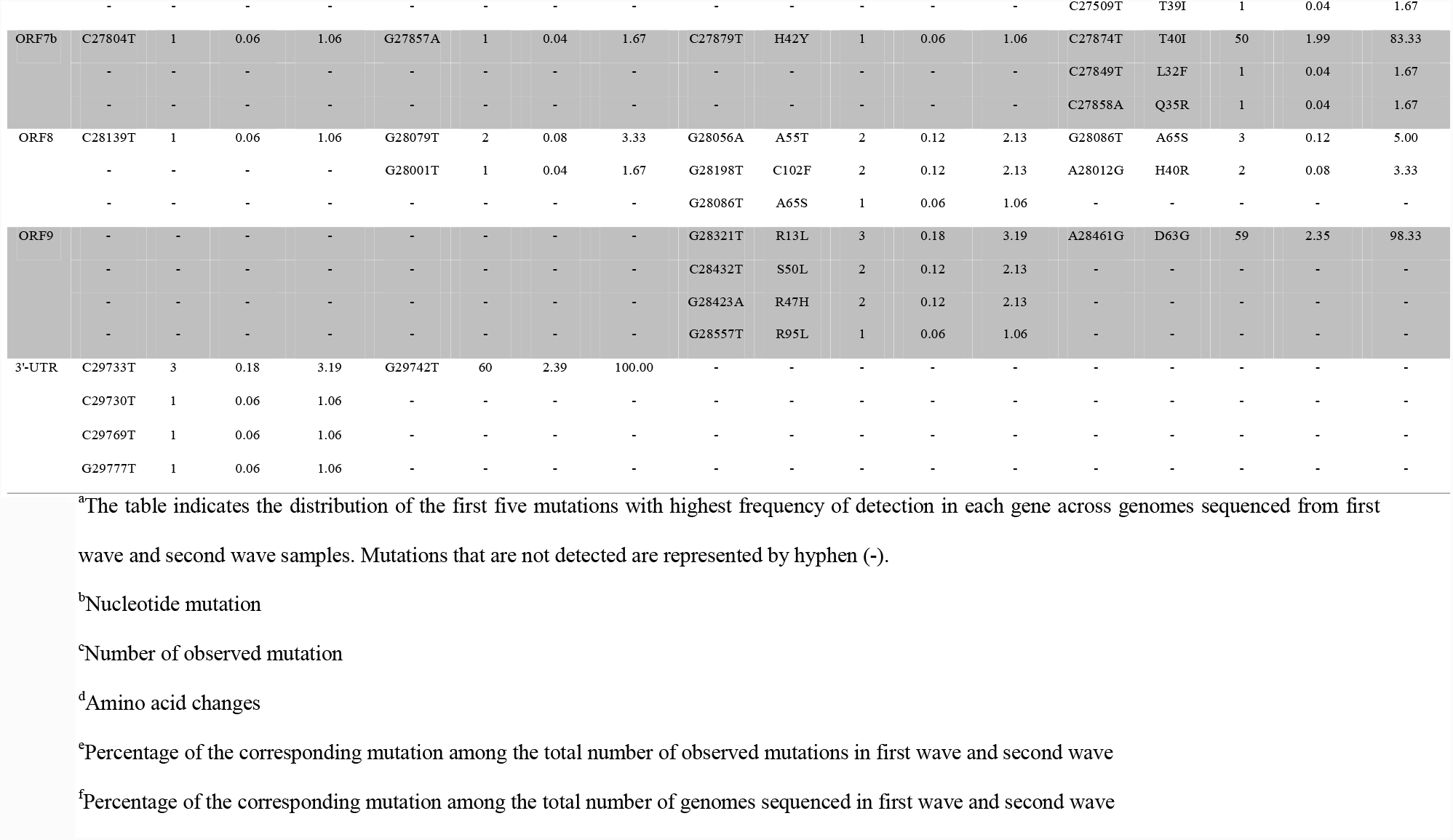
Top five mutation detected in the genes across the sequenced SARS-CoV-2 genomes^a^.

The NE India reported numerous incidences of VOCs of SARS-CoV-2, especially Delta and AY sublineages (Figure 3 and Figure 4). VOCs emerged in the region as early as December 2020, and Delta variant was first detected in January 2021 in Assam (Figure 4). The data showed the displacement of normal variants in first wave by the VOCs in the second wave, which was completely dominated by Delta and AY sublineages, indicating intra-lineage mutation.

**Figure 3.**
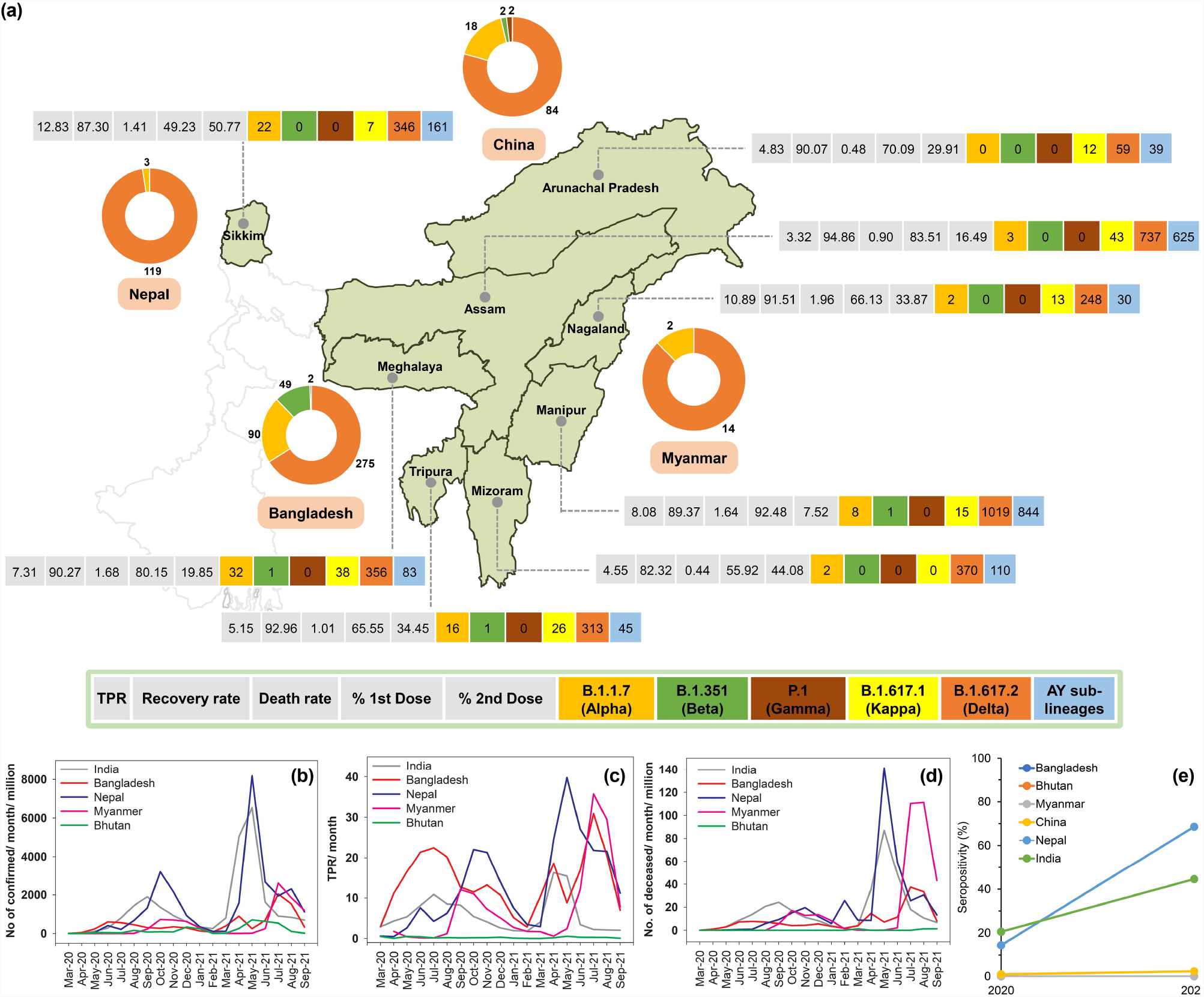
Distribution of variants of SARS-CoV-2 in various states of North East region of India and in the neighboring countries sharing international border with the region (a) comparison with confirmed COVID-19 cases (b), test positivity rate (TPR) (c), deaths (d), and seropositivity rate (e). (a) Demographics of qRT-PCR TPR, recovery rate, death rate, and vaccination rate are also given for the states of North East India. Data on the variants of SARS-CoV-2 indicate the cumulative total of each variant detected since the first sample from the state and the country was sequenced. The genome data for the North East states of India were retrieved from the Indian COVID-19 Genome Surveillance data hub of the Indian SARS-CoV-2 Genomics Consortium (INSACOG) (http://clingen.igib.res.in/covid19genomes/), and the EpiCoV database in GISAID server (https://www.gisaid.org/) (retrieved on 15 October 2021). The genome dataset for the state of Assam and Arunachal Pradesh also included 719 and 96 genome sequences, respectively, that were generated in the present study. Month-wise evolution of the variants in the states are given in Figure 4. The variant data for the neighboring countries of Nepal, Bhutan, China, Myanmar and Bangladesh were obtained from the global SARS-CoV-2 genome dataset available in EpiCoV database in GISAID server (https://www.gisaid.org/) (retrieved on 15 September 2021). Only the variants of concern and variant of interest detected in the states of North East India are shown. The variants of concern are B.1.1.7 (Alpha), B.1.351 (Beta), P.1 (Gamma), and B.1.617.2 (Delta), and AY sublineages, and B.1.617.1 (Kappa) is the variant of interest. The data related to COVID-19 test, recovery and death are represented by TPR, recovery rate and death rate, respectively, as percentage value, and are calculated using the data retrieved from www.covid19india.org (retrieved on 17 July 2021) and include data collected from April 2020 – July 2021. The TPR, recovery rate and death rate represent the cumulative of both first wave and second wave. The TPR is calculated irrespective of the type of method used for testing. The data related to vaccination is represented by percentage of the first and second doses administered during 16 January 2021 to 16 July 2021, irrespective of the type of vaccine, age, gender, vaccination session and vaccination sites. The vaccination data was retrieved from www.covid19india.org (retrieved on 17 July 2021) and validated with the data available in the official website of Ministry of Health and Family Welfare (MoHFW), Govt. of India (www.mohfw.gov.in). (b, c, d) Data for the graphs depicting COVID-19 confirmed cases per million individuals, TPR, and number of deaths per million individuals in the neighboring countries sharing international border with North East region of India was obtained from Our World in Data (https://ourworldindata.org/coronavirus) from 1 March 2020 – 30 September 2021. (e) The graph represents the seropositivity rate of India and neighboring countries. Seropositivity rate in both 2020 and 2021 for India, China and Nepal were represented as the average of three different time points of each year. Rate of seropositivity data is not available for Bangladesh, Bhutan and Myanmar. Data was obtained from Serotracker as on 24 October 2021. (https://serotracker.com/en/Explore)

**Figure 4.**
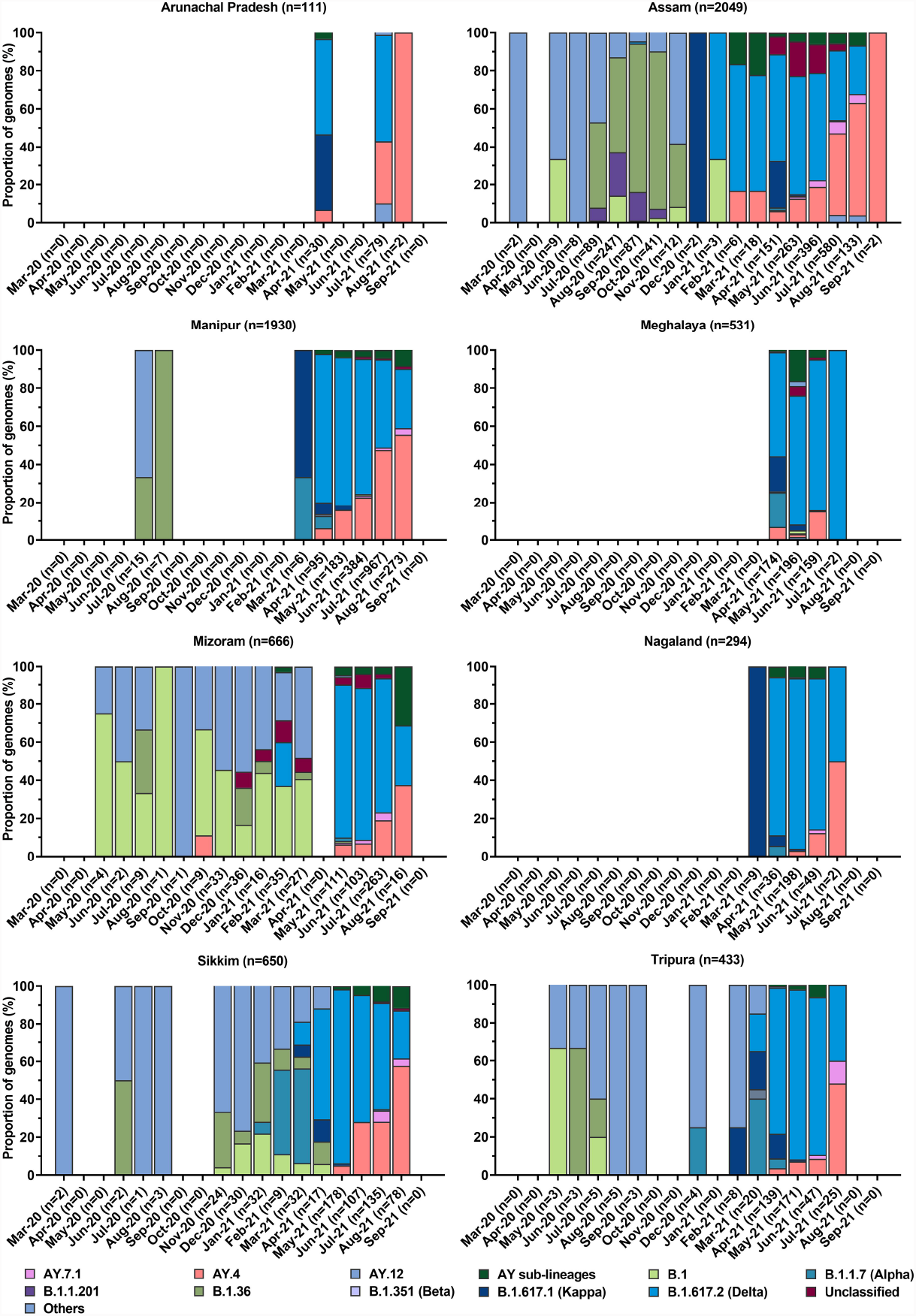
Evolution of SARS-CoV-2 variants at different time points of their detection in the states of North East India. The data shows the displacement of normal variants in first wave by the variant of interest and variants of concern in the second wave. The second wave was completely dominated with Delta variant and its AY sublineages, indicating intra-lineage mutation and arrest of emergence of new variants. The genome data were retrieved from the Indian COVID-19 Genome Surveillance data hub of the Indian SARS-CoV-2 Genomics Consortium (INSACOG) (http://clingen.igib.res.in/covid19genomes/), and the EpiCoV database in GISAID server (https://www.gisaid.org/) (retrieved on 15 October 2021). The genome dataset for the state of Assam and Arunachal Pradesh also included 719 and 96 genome sequences, respectively, that were generated in the present study.

### Serological study of cohort pre-and post-vaccination

In the cohort (*n*=35), pan-IgG-N antibody increased from 8.57% in August 2020 to 28.57%, 31.42%, 45.71% and 45.71% in February 2021, June 2021, July 2021 and August 2021, respectively, correlating well with the increase in COVID-19 incidences (Table 3). Three subjects tested seropositive in 10^th^ August 2020, which continuously persisted till 14^th^ August 2021 (Supplementary Figure S4) highlighting the persistence of antibody against SARS-CoV-2 for more than a year in some individuals. The neutralizing activity gradually increased from August 2020 onwards and reached 88.57% by mid July 2021 (Table 3). There was a 2.33-fold increase in neutralizing activity in February 2020 compared to August 2020. However, neutralizing activity increased by 1.57-fold in June 2021 and more than 4-fold in July 2021 compared to February 2021. Interestingly, IgG-N antibody remained low compared to neutralizing activity after February 2020. The increase in neutralizing activity within a short span of time could be due to the effect of vaccination (Supplementary Table S6). On the other hand, only 28.57% (n=10) acquired COVID-19 infection (5 subjects during first wave and 5 subjects during second wave) in the entire study. However, prevalence of neutralizing antibody started to decrease from last week of July 2021 reaching as low as 68.57% by the end of July 2021.Vaccine breakthrough was observed in only one single individual vaccinated with Covishield who was asymptomatic seropositive during first wave. In this cohort group, three individuals were diabetic among them only one contracted COVID-19 post single-dose vaccination during second wave. Additionally, three out of five hypertensive individuals developed COVID-19 infection. None among the 19 individuals with smoking habits were reported to be infected with COVID-19. Out of the 19 alcoholic and 17 tobacco-consuming individuals, only 2 cases with both the habits acquired COVID-19 infection. Moreover, it was observed that individuals infected with COVID-19 prior to vaccination showed higher neutralizing activity irrespective of lifestyle and co-morbidity.

**Table 3:** IgG-N reactive antibody and neutralizing activity against SARS-CoV-2 among the cohort of 35 people^a^.

| Parameters | 2020<br>(n = 35) | 2021<br>(n = 35) |  |  |  |  |  |  |
| --- | --- | --- | --- | --- | --- | --- | --- | --- |
| Sampling Date | 10 August | 10 February | 10 June | 10 July | 16 July | 24 July | 31 July | 14 August |
| **IgG-N antibody [n (%)]** | 3 (8.57%) | 10 (28.57%) | 11 (31.42%) | 16 (45.71%) | 16 (45.71%) | 16 (45.71%) | 16 (45.71%) | 16 (45.71%) |
| **Neutralizing antibody [n (%)]** | 3 (8.57%) | 7 (20%) | 11 (31.42%) | 29 (82.85%) | 31 (88.57) | 25 (71.41%) | 24 (68.57%) | 26 (74.28%) |
| **Vaccinated, Single dose [n (%)]** | - | - | 20 (57.1%) | 15 (42.85%) | - | - | - | - |
| **Vaccinated, Double dose [n (%)]** | - | - | 10 (28.57%) | 4 (11.42%) | - | - | - | - |
| **qRT-PCR positive [n (%)]** | 5 (14.2%) | - | 1 (2.85%) | 1 (2.85%) | 1 (2.85%) | - | - | 2 (5.71%) |
| **Vaccine breakthrough<sup>b</sup> [n (%)]** | - | - | - | - | - | - | - | 1 (2.85%) |

<sup>a</sup>Cohort group includes 29 male and 6 female participants aged between 25 to 55 years with an average age of 35.77 years.
<sup>b</sup>Vaccine breakthrough is defined as one who got infected after receiving both doses and completed at least 14 days of follow-up after the second dose.

## Discussion

North East India is a unique part of India with significant geographical and inhabitant population variation. Considering this unique regional importance and as part of global initiative to generate and exchange genomic and serological profile of SARS-CoV-2 transmission and evolution, we have conducted this study of NE India.

Our meta-analysis of incidences of COVID-19 in NE India in comparison to trend observed in overall Indian population showed that number of incidences across NE state including Assam is relatively low (Figure 1). However, it is not known whether the inherent immune-genetic factor or epigenetic factors of the unique host population had played any role in preventing or augmenting the incidences. Conversely, the higher rate of incidences in overall Indian population in comparison to NE Indian states can be due to extremely higher local incidences in some part of India such as Maharashtra, Delhi, Karnataka. Subsequently, this was followed by rapid rise in the incidences leading to the peak of second wave with highest incidences during May-June 2021 (Figure 1). Interestingly, NE states of India showed much sharper rise in incidences. When we normalized the data per 1000 cases and carried out analysis of incidences of COVID-19 among the different states of NE India, we observed interesting variations in number of waves and peak intensities between the states (Supplementary Table S7 & Table S8). During first wave, the increase in infection was steady and gradual across the NE India, which might have led to the emergence of various variants while the virus was transmitted from person to person and adapt with the new host. Awareness, self-precaution, physical and social distancing, and strict lockdown measures prescribed by the state governments might have resulted to the rapid and effective control of the COVID-19 transmission in the region during first wave on the basis of the skewed distribution observed during first wave. The observed rapid rise in infection and community transmission in NE India during the second wave might have resulted from the emergence of VOCs B.1.617.2 and AY sublineages, with high transmission and immune escape properties [9]. This was further elevated with relaxation in preventive COVID-19 measures by the respective state governments.

In our serological study, it was observed that the prevalence of seropositivity increased while the frequency of neutralizing antibody decreased both in the intermittent phase and in second wave compared to those seen in first wave. The seropositivity rate was observed to be 10.61% during first wave, which supported our earlier finding [10], while the seropositivity rate during second wave increased nearly 4-fold. Additionally, the seropositivity in the second wave was also higher to those seen in the intermittent phase, which might be due to the emergence of new variants such as B.1.617.2 and AY sublineages. Several explanations have put forwarded as possible factors for generation of multi-waves and their characteristics in major pandemics. Some of these are co-infection [11], pandemic fatigue [12], sub-optimal prevention and control measures during intermittent endemic period [13], climatic variations [14] and genetic drift of the infectious agent associated with diffusion and herd immunity [15, 16]. Moreover, the increase in seropositivity and neutralizing antibody in older subjects unlike younger in the second wave may suggest the influence of age-selective mass vaccination programme in addition to possible emergence of new mutants. In the cohort group study, it was recorded that anti-SARS-CoV-2 antibody could last more than a year, which was also reported by Xiao et al., 2021 implying that some individuals have the capacity to sustain antibodies for a relatively longer period of time for prolonged protection against SARS-CoV-2 [17]. Additionally, it was observed that individuals infected with COVID-19 prior to single or double dose vaccination resulted in higher neutralizing activity irrespective of lifestyle and health condition. This result corroborated by the work of Reynolds et al. 2021 where they reported that a single dose of Pfizer/BioNTech mRNA vaccine can enhance higher T-cell immunity and antibody secreting memory B-cell response, and neutralizing antibodies against B.1.1.7 and B.1.351 variants in individuals with prior COVID-19 infection [18]. The observance of lower neutralizing antibody in second wave as compared to first wave among the individuals was possibly due to reduced sensitivity of SARS-CoV-2 variant B.1.617.2 to antibody neutralization as also reported by other groups [19]. In addition, neutralizing antibodies decreased in the vaccinated 35 cohort group after few months, which could be assumed as possible shifting of humoral immunity of antibodies towards generation of memory B cells for longer uses [20, 21].

The whole genome sequences allowed us to detect the representative diversity of SARS-CoV-2 viruses in NE India, and compared with other states in the NE India. Most importantly, our genome sequencing work sought to confirm the first occurrence of VOC in NE India. It is to be noted here that B.1.617.2 was first identified in India in August 2020, and our findings confirmed the earliest occurrence of B.1.617.2 in the NE state of Assam within one month period in September 2020. This clearly indicated the fact that the transmission of this highly contagious B.1.617.2 variant was very quick and prompt from the mainland of India to NE India. Upon first occurrence of B.1.617.2 in Assam, a highly rapid transmission of this contagious variant and its various AY sublineages was observed in the NE India during second wave. We also observed that the occurrence of B.1.617.2 and AY sublineages was mostly associated with partially vaccinated population. This observation is in consistence with earlier reports [22, 23]. During May 2021, in the UK, it was reported that the secondary attack rate of B.1.617.2 was higher than B.1.1.7 in partially immunized population [24]. The linkage between the VOC and VOI were mainly characterized by unique substitution or deletion in the spike protein. Changes in L452R, E484Q and P681R amino acids were identified in Kappa variant while Delta was identified with changes in L452R and P681 R amino acids [25]. The changes in these three amino acid positions were directly linked with the strong binding affinity with the host as well as capability to escape from the neutralizing antibodies by SARS-CoV-2 [26]. Overall, the demonstration of genomic characterizations of SARS-CoV-2 variants of NE India increased the availability of geographical-based genomes of SARS-CoV-2 variants and thereof illustrated emerging trends in variations in the genomes of SARS-CoV-2 variants.

During the second wave while India had been witnessing the massive burden of rapid surge in COVID-19 cases, most of the neighboring countries sharing international border with NE region of India had relatively small number of cases yet (Figure 3). However, Bhutan was an exception, where it was experiencing one of its largest surges even though 62% of the population had received at least one dose of a vaccine [27]. The surge in cases in Bangladesh corresponded with widespread detection of variant B.1.351 (Beta) in February and March 2021 during the second wave [28], which was later dominated by B.1.617.2. The surge in cases in India, particularly in its NE region, and Nepal corroborated with the increasing incidences of B.1.617.2. The surge is cases in Bhutan and Myanmar, though started after India and its NE region reported rapid decline in the cases, coincided with the prevalence of B.1.617.2. Overall, it is indicative that the second wave in the neighboring countries sharing international border with NE region of India followed similar pattern of rise and fall of cases and is driven by the B.1.617.2 variant. Considering the fragile and possible porous nature of these international borders, it is highly likely that undetected cross-border spillage of SARS-CoV-2 variants, particularly B.1.617.2 may have occurred during the second wave.

## Conclusion

The present study demonstrates that although qRT-PCR testing shows correlation with serological testing, each approach can derive different epidemiological profile of evolution and transmission of SARS-Cov-2 in a given population. First wave of COVID-19 in NE India witnessed low transmission with more diverse evolution of SARS-CoV-2 variants, which might have prepared the virus to develop higher transmissibility, more resilience to host immune response through acquired immune escape potential and more lethal virulence to drive the second wave. The widely varied prevalence of SARS-CoV-2 variants across the NE India implies potential of spread of any communicable diseases through and beyond this region. Detection of only one dominating variant of concern throughout second wave in Assam is also indicative of stringent tracking, tracing and treating policies undertaken by the local state government. Vaccination has a strong role to play in reducing the infection, stabilizing the immunity against recurrent infection and thus possibly breaking the chain for further spread. Additional studies are warranted to understand how the virus will evolve post vaccination, which will dictate the outcome of any possible future wave of COVID-19.

## Data Availability

Data that have been taken from open source databases are readily available, and if needed all data produced in the present study are available upon reasonable request to the authors.

## Conflicts of interest

The authors declare no conflicts of interest related to this article.

## Funding

This work was supported by the Council of Scientific and Industrial Research (CSIR), Government of India under its intramural COVID-19 projects entitled “Testing and Management of COVID-19” [grant number MLP0158] and “Genome Sequencing of COVID-19” [grant number MLP1019].

## Role of the funding source

The funding source had no role in study design, data collection, data analysis, data interpretation, or writing of the manuscript.

## Acknowledgments

The authors would like to acknowledge all the doctors, technical staff, and nurses of the Clinical Centre, CSIR-NEIST, Jorhat, and CSIR-NEIST COVID-19 Warriors of the institute for their support in organizing camps for blood sample collection and vaccination. The Indian SARS-CoV-2 Genomics Consortium (INSACOG) and its Scientific and Clinical Advisory Group (SCAG) are thanked for the constant support in establishing the Genome Sequencing Facility in CSIR-NEIST, Jorhat and in subsequent genome sequencing of SARS-CoV-2 variants. The authors thank Dr Anurag Agrawal, Director, CSIR-Institute of Genomics and Integrative Biology, New Delhi, India for providing critical comments for improving the manuscript.

## Authors’ contributions

GNS and JK have conceptualized the study. RW, PB, TP and PM along with GNS and JK involved in its design, project administration, management of resources, supervision, funding acquisition, validation, investigation, critical inputs, and preparing the first draft and critical revising of the manuscript. GNS and JK were the coordinators of the COVID-19 projects and led the relationship with IDSP, Department of Health and Family Welfare, Government of Assam and connection amongst the group of scientists. RW, PB, PM, TP and RK were responsible for leading the study execution. NS, HJM, HS, AKS, NV, CC, RKS, PD, GG, YBT, MGS and SBW were involved in literature search, meta-data collection, data acquisition, data curation, investigation, methodology, formal analysis, compiling and collating figures and tables, data interpretation. RK, RW, PB, PM, TP, MGS, GG and YBT were responsible for testing of COVID-19 samples, result generation and verification of COVID-19 qRT-PCR test data. PM, TP, RKS and PD conducted the serological investigations, data acquisition thereof and its interpretation. RW, PB and MGS carried out library preparation, loading and running of genome sequencing, and sequence data analysis. GG, YBT, MGS, and RKS contributed in literature search, meta-data collection and data acquisition, and also in organizing camps for enrolment of study participants, blood sample collection, and logistics required for serological study. JK and GNS contributed in data interpretation, data analysis, manuscript reviewing and editing. All authors had full access to all the data in the study and were responsible for the final decision to submit the manuscript for publication.

## Notes

### Competing Interest Statement

The authors have declared no competing interest.

### Author Declarations

Institutional Human Ethics Committee of CSIR-NEIST gave ethical approval for the present work vide approval no. IHEC/NEIST20-21/201.

