## Supplementary Table for "Seroepidemiological and genomic investigation of COVID-19 spread in North East region of India"

**Supplementary Appendix 1**

**(Supplementary tables)**

**Table S1: Test positivity rate of COVID-19 in Jorhat, Assam during first wave (June-September 2020) and second wave (May-July, 2021)^a^**

|  | **First wave** | | **Second wave** | |
| --- | --- | --- | --- | --- |
| **Parameters** | **Samples tested** | **Positive case**  **(TPR %)** | **Samples tested** | **Positive case**  **(TPR %)** |
| Total | 12879 | 571 (4.43) | 3416 | 248 (7.26) |
| **Gender** | | | | |
| Male | 8765 | 379 (4.32) | 2110 | 154 (7.30) |
| Female | 4097 | 192 (4.68) | 1306 | 94 (7.20) |
| Not disclosed | 17 | - | - | - |
| **Age groups** | | | | |
| ≤10 | 381 | 10 (2.62) | 74 | 7 (9.46) |
| 11-30 | 4804 | 210 (4.37) | 1098 | 72 (6.56) |
| 31-60 | 7453 | 338 (4.53) | 2053 | 154 (7.50) |
| ≥61 | 241 | 13 (5.39) | 191 | 15 (7.80) |

^a^Data represents the RT-PCR test positivity rate of the oro- and naso-pharyngeal swab samples collected during first wave (June-September 2020) and second wave (May-July, 2021) at COVID-19 Testing Lab, CSIR-NEIST, Jorhat, Assam.

**Table S2: Infectivity of the participants and their positivity in serological and neutralizing assays for SARS-CoV-2 in** **August, 2020**

| **Parameter** | **Participants tested, n** | **COVID-19 positive** | **Unweighted positivity, % (95% CI)** | **IgG-N** | **Unweighted positivity, % (95% CI)** | **IgG-S** | **Unweighted positivity, % (95% CI)** | **IgM-S** | **Unweighted positivity, % (95% CI)** | **Neutralizing antibodies** | **Unweighted positivity, % (95% CI)** |
| --- | --- | --- | --- | --- | --- | --- | --- | --- | --- | --- | --- |
| **Overall** | 724 | 33 | 4.56%  (3.04-6.07) | 77 | 10.61% (8.37-12.86) | 64 | 8.84% (6.77-10.91) | 7 | 0.97%  (0.25-1.68) | 70 | 9.67%  (7.51-11.82) |
| **Gender** | | | | | | | | | | | |
| Male | 521 | 25 | 4.8%  (2.96-6.63) | 60 | 11.49%  (8.75-14.23) | 53 | 10.17  (7.57-12.77) | 7 | 1.34 %  (0.35-2.33) | 54 | 10.36%  (7.74-12.98) |
| Female | 203 | 8 | 3.94%  (1.26-6.61) | 17 | 8.36%  (4.55-12.16) | 11 | 5.42%  (2.30-8.53) | 0 | 0 | 16 | 7.88%  (4.17-11.58) |
| **Age group** | | | | | | | | | | | |
| <30 | 179 | 2 | 1.12%  (-0.422-2.65) | 19 | 10.59%  (6.08-15.01) | 15 | 8.38%  (4.32-12.44) | 0 | 0 | 18 | 10.05%  (5.65-14.46) |
| 30-60 | 453 | 27 | 5.96%  (3.78-8.14) | 52 | 11.45%  (8.52 | 43 | 9.49%  (6.79-12.19) | 7 | 1.54%  (0.41-2.68) | 45 | 9.93%  (7.17-12.68) |
| >60 | 92 | 4 | 4.35%  (0.18-8.51) | 6 | 6.51%  (1.46-11.55) | 6 | 6.52%  (1.47-11.56) | 0 | 0 | 7 | 7.61%  (2.19-13.02) |

**Table S3: Infectivity of the participants and their positivity in serological and neutralizing assays for SARS-CoV-2 in** **February, 2021**

| **Parameter** | **Participants tested, n** | **COVID-19 positive** | **Unweighted positivity, % (95% CI)** | **IgG-N** | **Unweighted positivity, % (95% CI)** | **IgG-S** | **Unweighted positivity, % (95% CI)** | **IgM-S** | **Unweighted positivity, % (95% CI)** | **Neutralizing antibodies** | **Unweighted positivity, % (95% CI)** |
| --- | --- | --- | --- | --- | --- | --- | --- | --- | --- | --- | --- |
| **Overall** | 866 | 58 | 6.69%  (5.03-8.36) | 349 | 40.21% (36.95-43.48) | 146 | 16.86%  (14.36-19.35) | 31 | 3.57%  (2.34-4.81) | 182 | 21.01%  (18.30-23.72) |
| **Gender** | | | | | | | | | | | |
| Male | 613 | 44 | 7.17%  (5.13-9.22) | 258 | 42%  (38.09-45.91) | 110 | 17.94%  (14.90-20.98) | 26 | 4.24%  (2.64-5.83) | 135 | 22.02%  (18.74-25.30) |
| Female | 253 | 14 | 5.53%  (2.71-8.35) | 91 | 35.89%  (29.98-41.80) | 36 | 14.23%  (9.92-18.53) | 5 | 1.97%  (0.26-3.69) | 47 | 18.57%  (13.78-23.36) |
| **Age group** | | | | | | | | | | | |
| <30 | 306 | 7 | 2.29%  (0.61-3.96) | 127 | 41.42% (35.90-46.94)) | 54 | 17.65%  (13.37-21.92) | 7 | 2.28%  (0.61-3.96) | 68 | 22.22%  (17.56-26.88) |
| 30-60 | 493 | 43 | 8.72%  (6.23-11.21) | 194 | 39.27%  (34.96-43.58) | 83 | 16.83%  (13.51-20.14) | 23 | 4.66%  (2.80-6.53) | 105 | 21.29%  (17.68-24.91) |
| >60 | 67 | 8 | 11.94%  (4.17-19.7) | 28 | 41.71%  (29.90-53.51) | 9 | 13.43%  (5.26-21.59) | 1 | 1.49%  (-1.41-4.39) | 9 | 13.43%  (5.26-21.59) |

**Table S4: Infectivity of the participants and their positivity in serological and neutralizing assays for SARS-CoV-2 in** **June, 2021**

| **Parameter** | **Participants tested, n** | **COVID-19 positive** | **Unweighted positivity,**  **% (95% CI)** | **IgG-N** | **Unweighted positivity,**  **% (95% CI)** | **IgG-S** | **Unweighted positivity,**  **% (95% CI)** | **IgM-S** | **Unweighted positivity,**  **% (95% CI)** | **Neutralizing antibodies** | **Unweighted positivity,**  **% (95% CI)** |
| --- | --- | --- | --- | --- | --- | --- | --- | --- | --- | --- | --- |
| **Overall** | 436 | 34 | 7.79%  (5.28-10.31) | 202 | 46.23%  (41.55-50.91) | 69 | 15.82%  (12.39-19.25) | 16 | 3.66%  (1.90-5.43) | 140 | 32.11% (27.72-36.49) |
| **Gender** | | | | | | | | | | | |
| Male | 314 | 28 | 8.91%  (5.76-12.07) | 180 | 57.21%  (51.73-62.68) | 55 | 17.51%  (13.31-21.72) | 12 | 3.82%  (1.70-5.94) | 125 | 39.81% (34.39-45.22) |
| Female | 122 | 6 | 4.92%  (1.08-8.75) | 22 | 17.99%  (11.17-24.81) | 14 | 11.47%  (5.81-17.13) | 4 | 3.27%  (0.11-6.43) | 15 | 12.29%  (6.46-18.12) |
| **Age group** | | | | | | | | | | | |
| <30 | 121 | 10 | 8.26%  (3.35-13.17) | 29 | 23.91%  (16.31-31.52) | 19 | 15.70%  (9.21-22.18) | 4 | 3.30%  (0.12-6.41) | 35 | 28.92% (20.84-37) |
| 30-60 | 287 | 20 | 6.97%  (4.02-9.91) | 155 | 53.89%  (48.13-59.66) | 41 | 14.28%  (10.23-18.33) | 9 | 3.13%  (1.11-5.15) | 86 | 29.96% (24.66-35.26) |
| >60 | 28 | 4 | 14.28%  \(1.32-27.24) | 18 | 64.15%  (46.39-81.91) | 9 | 32.14%  (14.84-49.44) | 3 | 10.71%  (-0.74-22.17) | 19 | 57.85% (50.55-85.15) |

**Table S6: Details of vaccination and COVID-19 positivity of the 35 cohort group**

| **Subject** | **Gender** | **Age group** | **Dose 1_Vaccine type** | **Dose 2_Vaccine type** | **RT-PCR result**  **(First wave)** | **RT-PCR result (Second wave)** | **Vaccine breakthrough infection** |
| --- | --- | --- | --- | --- | --- | --- | --- |
| Subject 1 | Male | ≤30 | Covishield | - | Negative | Negative | No |
| Subject 2 | Male | 41-50 | Covaxin | - | Negative | Negative | No |
| Subject 3 | Male | 41-50 | Covishield | - | Negative | Negative | No |
| Subject 4 | Female | 31-40 | Covishield | - | Negative | Negative | No |
| Subject 5 | Male | 31-40 | Covishield | - | Negative | Positive | No |
| Subject 6 | Male | 41-50 | Covishield | Covishield | Negative | Negative | No |
| Subject 7 | Male | 41-50 | Covishield | - | Negative | Negative | No |
| Subject 8 | Male | ≤30 | Covishield | - | Negative | Positive | No |
| Subject 9 | Male | 41-50 | Covaxin | - | Negative | Negative | No |
| Subject 10 | Male | 51-60 | Covishield | Covishield | Negative | Negative | No |
| Subject 11 | Male | ≤30 | Covishield | - | Negative | Negative | No |
| Subject 12 | Male | 31-40 | Covishield | Covishield | Negative | Negative | No |
| Subject 13 | Male | 41-50 | Covaxin | Covaxin | Negative | Negative | No |
| Subject 14 | Male | 31-40 | Covishield | - | Positive | Negative | No |
| Subject 15 | Male | 31-40 | Covishield | - | Negative | Negative | No |
| Subject 16 | Male | 41-50 | Covishield | - | Positive | Negative | No |
| Subject 17 | Male | 31-40 | Covishield | Covishield | Negative | Negative | No |
| Subject 18 | Male | 31-40 | Covishield | Covishield | Positive | Negative | No |
| Subject 19 | Female | ≤30 | Covishield | - | Negative | Negative | No |
| Subject 20 | Male | 51-60 | Covishield | Covishield | Negative | Negative | No |
| Subject 21 | Female | ≤30 | Covishield | - | Negative | Positive | No |
| Subject 22 | Female | ≤30 | Covishield | - | Negative | Negative | No |
| Subject 23 | Male | 31-40 | Covishield | Covishield | Negative | Negative | No |
| Subject 24 | Male | 41-50 | Covishield | Covishield | Negative | Negative | No |
| Subject 25 | Male | ≤30 | Covishield | - | Positive | Negative | No |
| Subject 26 | Female | ≤30 | Covishield | - | Negative | Positive | No |
| Subject 27 | Male | ≤30 | Covishield | - | Negative | Negative | No |
| Subject 28 | Male | 31-40 | Covishield | Covishield | Negative | Positive | Yes |
| Subject 29 | Male | ≤30 | Covishield | - | Negative | Negative | No |
| Subject 30 | Male | 31-40 | Covaxin | Covaxin | Positive | Negative | No |
| Subject 31 | Male | 41-50 | Covaxin | - | Negative | Positive | No |
| Subject 32 | Male | 31-40 | Covaxin | Covaxin | Negative | Negative | No |
| Subject 33 | Male | 31-40 | Covishield | Covishield | Negative | Negative | No |
| Subject 34 | Male | 31-40 | Covishield | Covishield | Negative | Negative | No |
| Subject 35 | Female | ≤30 | Covishield | - | Negative | Negative | No |

**Table S7: Quantitative analysis of peaks and gaps in terms of confirmed cases in North Eastern region of India**

| **State** | **No. of peaks** | **No. of waves** | **Gap^#^ between peaks** | **Intensity^@^ of peak 1** | **Duration^#^ of peak 1** | **Intensity^@^ of peak 2** | **Duration^#^ of peak 2** | **Skewed distribution of peak 1** | **Skewed distribution of peak 2** |
| --- | --- | --- | --- | --- | --- | --- | --- | --- | --- |
| Assam | 2 | 2 | 243 | 71770 | 334 | 158093 | 184 | L  (γ = 5.966) | R  (γ = 0.910) |
| Arunachal | 2 | 1 | 243 | 5684 | 334 | 8853 | 184 | L  (γ = 1.686) | R  (γ = 0.482) |
| Manipur | 2 | 1 | 212 | 7519 | 365 | 19165 | 153 | L  (γ = 2.406) | R  (γ = 0.544) |
| Meghalaya | 2 | 1 | 212 | 3813 | 365 | 18752 | 153 | L  (γ = 2.903) | R  (γ = 0.771) |
| Mizoram | 2 | 1 | 212 | 1103 | 365 | 7988 | 153 | L  (γ = 11.906) | L  (γ = 2.880) |
| Nagaland | 2 | 2 | 212 | 2884 | 334 | 7704 | 184 | L  (γ = 5.116) | L  (γ = 1.276) |
| Sikkim | 2 | 2 | 243 | 1279 | 334 | 7365 | 184 | L  (γ = 1.105) | R  (γ = 0.761) |
| Tripura | 2 | 2 | 243 | 14087 | 334 | 15964 | 184 | L  (γ = 4.064) | R  (γ = 0.534) |

A quantitative analysis of the peaks and gaps for all the states of north eastern region was performed based on the mean absolute deviation of the daily confirmed cases for the period Jun. 2020 to Jul. 2021. The gap between the peaks was measured in terms of ^#^number of days, intensity of the peaks in terms of ^@^number of confirmed cases and duration of a peak in terms of ^#^number of days. The skewed distribution was also performed as the ratio of left skewed distribution (AUC_L_) and right skewed distribution (AUC_R_) i. e.$\gamma=\frac{{AUC}_{L}}{{AUC}_{R}}$ **.** The left skewed distribution was the product of duration under the area in terms of days ($\propto_{L_{i}}$) and total cases in under the period ($\sigma_{L_{i}}$)**.** Similarly the right skewed distribution was computed. Mathematically, ${AUC}_{L}= \alpha_{L_{i}}\times\sigma_{L_{i}}$ & ${AUC}_{R}=\alpha_{R_{i}}\times\sigma_{R_{i}}$such that,

$\propto_{L_{i}}=\left| \frac{p_{i}-L_{i}}{L_{i+1}-L_{i}} \right|$	&	$\propto_{R_{i}}=\left| \frac{L_{i+1}-p_{i}}{L_{i+1}-L_{i}} \right|$, where *L_i_* is the point of start and end point for each peak and *p_i_* is the time point for the i^th^ peak.

$\sigma_{L_{i}}=\sum_{i=1}^{p_{i}} n_{i}$ & $\sigma_{R_{i}}=\sum_{i=p_{i+1}}^{L_{i+1}} n_{i}$ ,where n is the number of confirmed cases during α.

**Table S8: Quantitative analysis of peaks and gaps in terms of total deaths in North Eastern region of India**

| **State** | **No. of peaks** | **No. of waves** | **^#^Gap between peaks** | **^@^Intensity of Peak 1** | | **^#^Duration of Peak 1** | **^@^Intensity of Peak 2** | **^#^Duration of Peak 2** | **Skewed distribution of peak 1** | **Skewed distribution of peak 2** |
| --- | --- | --- | --- | --- | --- | --- | --- | --- | --- | --- |
| Assam | 2 | 2 | 243 | 391 | | 334 | 2058 | 153 | L  (γ = 2.138) | R  (γ = 0.962) |
| Arunachal | 2 | 1 | 92 | 21 | | 214 | 57 | 212 | L  (γ = 1.101) | L  (γ = 2.648) |
| Manipur | 2 | 1 | 273 | 113 | | 365 | 402 | 31 | L  (γ = 2.180) | R  (γ = 0.871) |
| Meghalaya | 2 | 1 | 212 | 39 | | 365 | 407 | 122 | L  (γ = 2.057) | R  (γ = 0.931) |
| Mizoram | 2 | 1 | 153 | 4 | | 182 | 53 | 122 | L  (γ = 3.279) | L  (γ = 8.276) |
| Nagaland | 2 | 2 | 212 | 27 | | 274 | 259 | 122 | R  (γ = 0.610) | L  (γ = 1.200) |
| Sikkim | 2 | 2 | 152 |  | 41 | 274 | 106 | 122 | L  (γ = 8.430) | L  (γ = 1.250) |
| Tripura | 2 | 2 | 173 |  | 171 | 273 | 164 | 153 | L  (γ = 1.941) | L  (γ = 4.058) |

All the calculations were performed using Python 3.7.0. A quantitative analysis of the peaks and gaps for all the states of north eastern region was performed based on the mean absolute deviation of the daily confirmed cases for the period Jun. 2020 to Jul. 2021. The gap between the peaks was measured in terms of ^#^number of days, intensity of the peaks in terms of ^@^number of confirmed cases and duration of a peak in terms of ^#^number of days. The skewed distribution was also performed as the ratio of left skewed distribution (AUC_L_) and right skewed distribution (AUC_R_) i. e.$\gamma=\frac{{AUC}_{L}}{{AUC}_{R}}$ **.** The left skewed distribution was the product of duration under the area in terms of days ($\propto_{L_{i}}$) and total cases in under the period ($\sigma_{L_{i}}$)**.** Similarly the right skewed distribution was computed. Mathematically, ${AUC}_{L}= \alpha_{L_{i}}\times\sigma_{L_{i}}$ & ${AUC}_{R}=\alpha_{R_{i}}\times\sigma_{R_{i}}$such that,

$\propto_{L_{i}}=\left| \frac{p_{i}-L_{i}}{L_{i+1}-L_{i}} \right|$	&	$\propto_{R_{i}}=\left| \frac{L_{i+1}-p_{i}}{L_{i+1}-L_{i}} \right|$, where *L_i_* is the point of start and end point for each peak and *p_i_* is the time point for the i^th^ peak.

$\sigma_{L_{i}}=\sum_{i=1}^{p_{i}} n_{i}$ & $\sigma_{R_{i}}=\sum_{i=p_{i+1}}^{L_{i+1}} n_{i}$ ,where n is the number of confirmed cases during α.
