## Supplementary Figure for "Seroepidemiological and genomic investigation of COVID-19 spread in North East region of India"

**Supplementary Appendix 2**

**(Supplementary figures)**

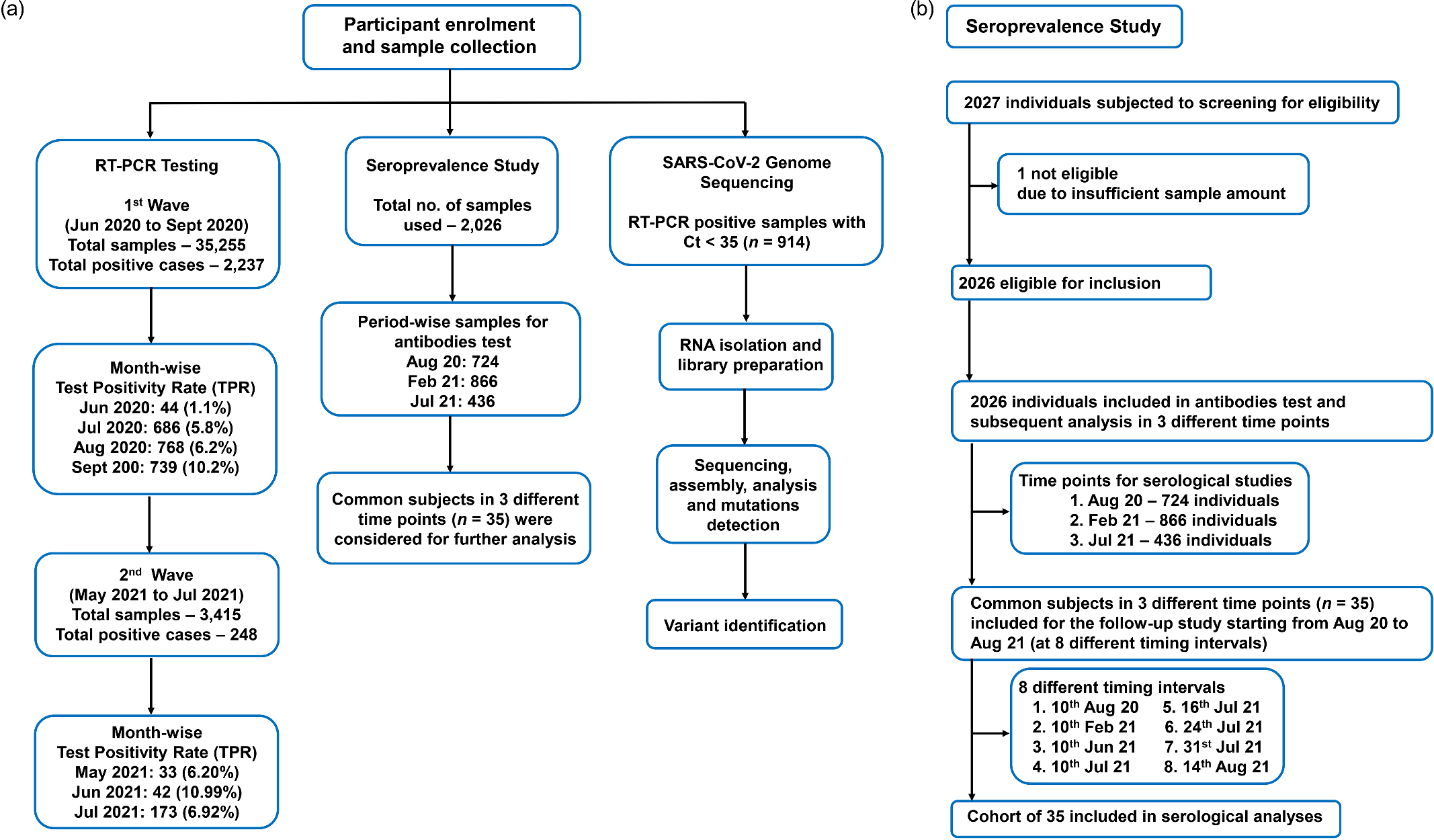

**Figure S1.** Schematic representation of the study approaches used in (a) enrolment of participants and (b) in the seroprevalence study

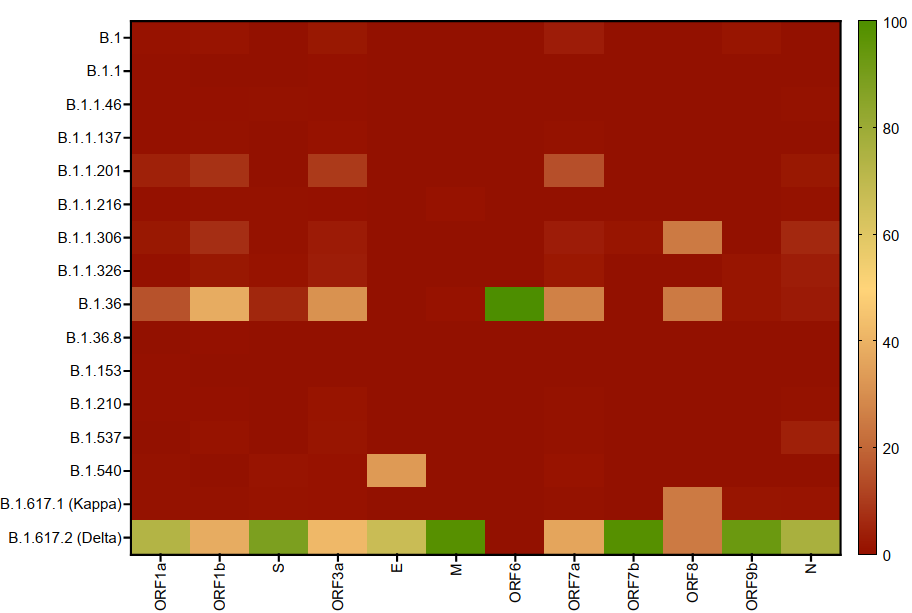

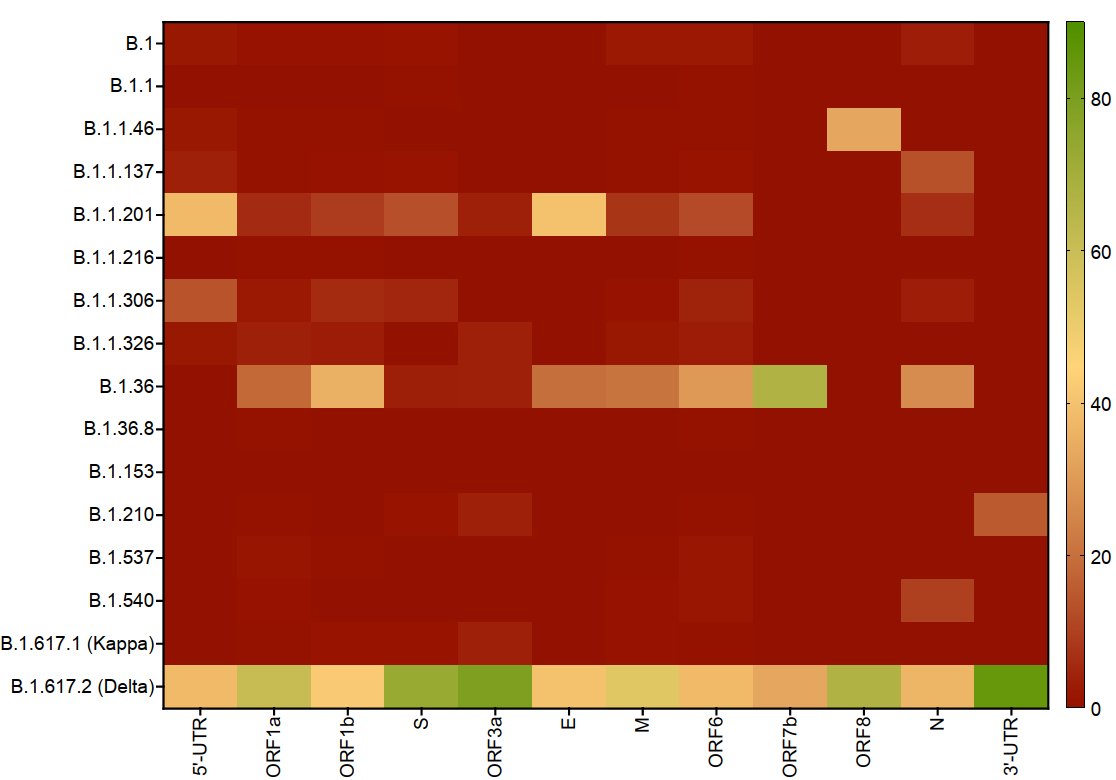

(a)	(b)

**Figure S2.** Heatmap showing the distribution of overall synonymous (a) and non-synonymous (b) substitution mutations in different variants of SARS-CoV-2 from first (July–August 2020) and second wave (May–June 2021) of North East India. Each data point indicates the normalized relative number synonymous (a) and non-synonymous (b) substitutions that are detected in each gene across the genomes analysed. Dark red designates detection of no mutation, yellow designates detection of lower number of mutations, and dark green designates highest number of mutations in the respective gene. The heatmaps were generated using GraphPad Prism 9 software.

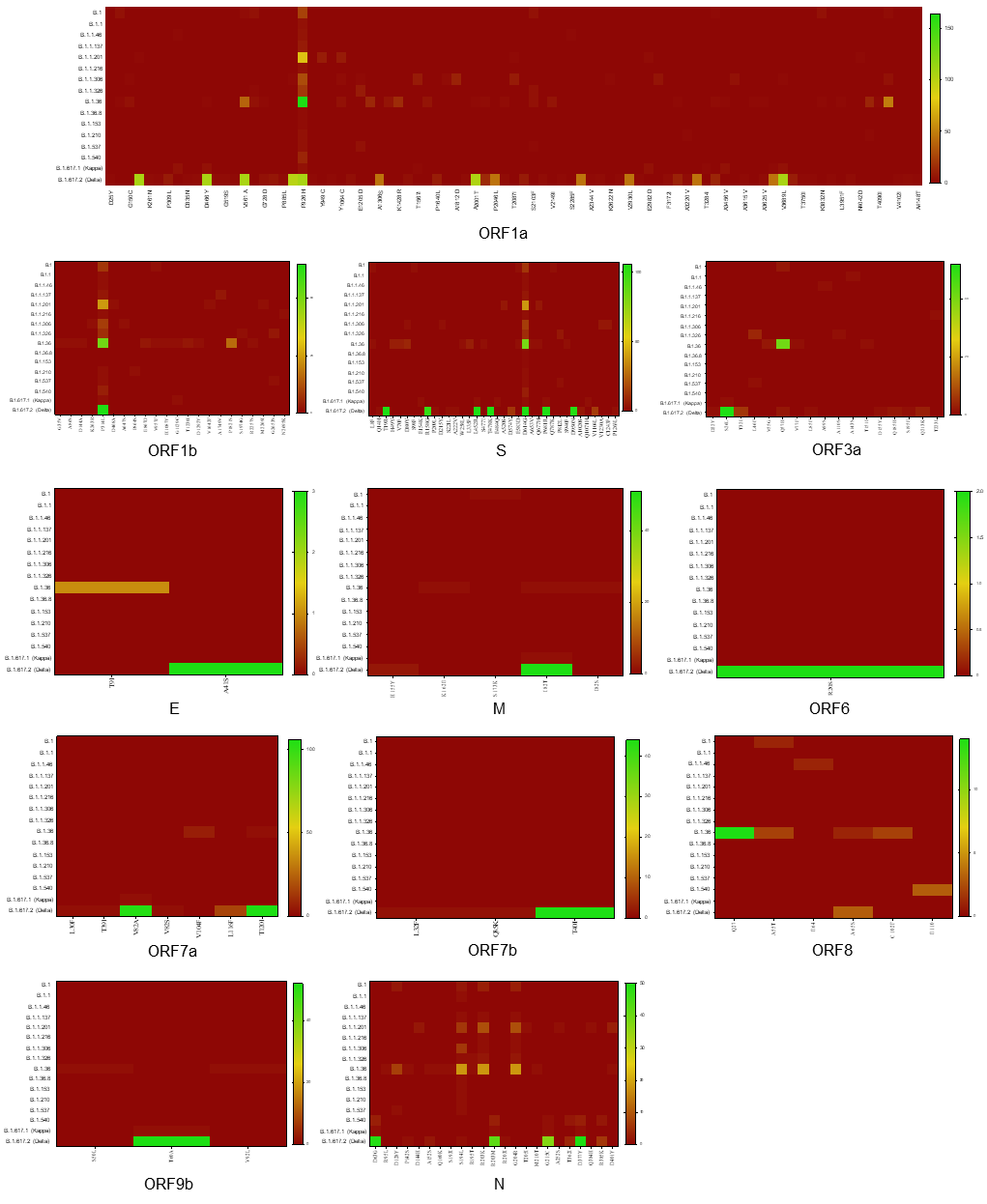

**Figure S3.** Heatmap showing the distribution of individual non-synonymous substitution mutations detected in the structural genes [spike (S), envelope (E), nucleocapsid (N) and membrane glycoprotein (M)] and non-structural genes (ORF1a, ORF1b, ORF3a, ORF6, ORF7a, ORF7b, ORF8 and ORF9b) in different variants of SARS-CoV-2 detected across the samples sequenced from first (July–August 2020) and second wave (May–June 2021) of North East India. Each data point indicates the cumulative number of non-synonymous substitutions that were detected in each gene across the genomes analysed. Dark red designates detection of no mutation, yellow designates detection of lower number of mutations, and dark green designates highest number of mutations in the respective gene. The heatmaps were generated using GraphPad Prism 9 software.

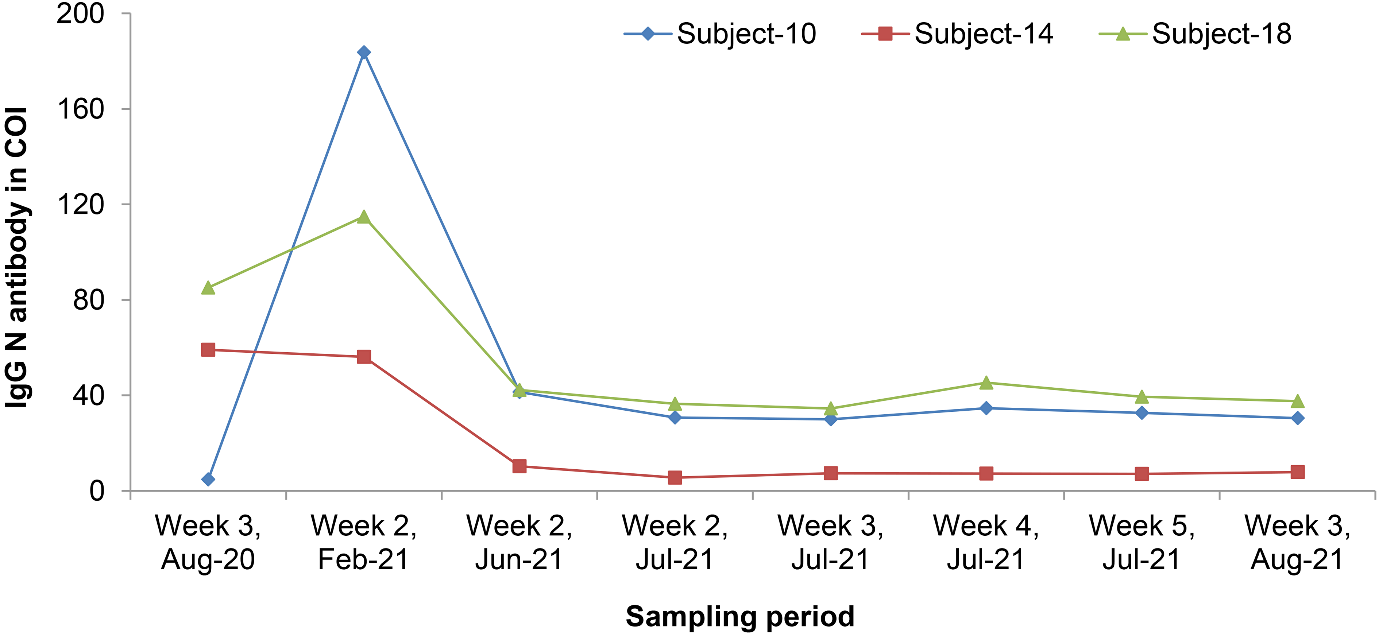

**Figure S4.** IgG-N antibody of three subjects throughout the study who were seropositive since August 2020**.**
